# How have mathematical models contributed to understanding the transmission and control of SARS-CoV-2 in healthcare settings? A systematic search and review

**DOI:** 10.1101/2023.03.17.23287403

**Authors:** David R M Smith, Sophie Chervet, Théo Pinettes, George Shirreff, Sofía Jijón, Ajmal Oodally, Kévin Jean, Lulla Opatowski, Solen Kernéis, Laura Temime

**Author notes:** Contributed equally (David R M Smith, Sophie Chervet). Contributed equally (Lulla Opatowski, Solen Kernéis, Laura Temime).

## Abstract

**Background:** Since the onset of the COVID-19 pandemic, mathematical models have been widely used to inform public health recommendations regarding COVID-19 control in healthcare settings.

**Objectives:** To systematically review SARS-CoV-2 transmission models in healthcare settings, and summarise their contributions to understanding nosocomial COVID-19.

**Methods:** Systematic search and review.

**Data sources:** Published articles indexed in PubMed.

**Study eligibility criteria:** Modelling studies describing dynamic inter-individual transmission of SARS-CoV-2 in healthcare settings, published by mid-February 2022.

**Participants and interventions:** Any population and intervention described by included models.

**Assessment of risk of bias:** Not appropriate for modelling studies.

**Methods of data synthesis:** Structured narrative review.

**Results:** Models have mostly focused on acute care and long-term care facilities in high-income countries. Models have quantified outbreak risk across different types of individuals and facilities, showing great variation across settings and pandemic periods. Regarding surveillance, routine testing – rather than symptom-based testing – was highlighted as essential for COVID-19 prevention due to high rates of silent transmission. Surveillance impacts were found to depend critically on testing frequency, diagnostic sensitivity, and turn-around time. Healthcare re-organization was also found to have large epidemiological impacts: beyond obvious benefits of isolating cases and limiting inter-individual contact, more complex strategies such as staggered staff scheduling and immune-based cohorting reduced infection risk. Finally, vaccination impact, while highly effective for limiting COVID-19 burden, varied substantially depending on assumed mechanistic impacts on infection acquisition, symptom onset and transmission. Studies were inconsistent regarding which individuals to prioritize for interventions, probably due to the high diversity of settings and populations investigated.

**Conclusions:** Modelling results form an extensive evidence base that may inform control strategies for future waves of SARS-CoV-2 and other viral respiratory pathogens. We propose new avenues for future models of healthcare-associated outbreaks, with the aim of enhancing their efficiency and contributions to decision-making.

## Introduction

SARS-CoV-2 transmission in healthcare settings has contributed significantly to the global health-economic burden of COVID-19. Healthcare settings are particularly vulnerable to COVID-19, due to dense concentrations of frail patients, high frequencies of at-risk medical procedures, and high rates of inter-individual contact. Both patients and healthcare workers (HCWs) have been at high risk of SARS-CoV-2 infection throughout the pandemic, resulting in major nosocomial outbreaks worldwide [1,2]. In England, for instance, an estimated 20% of patients hospitalized with COVID-19 before August 2020 acquired their infection in hospital [3], while 95,000-167,000 patients became infected during their hospital stay between June 2020 and March 2021 [4]. Further, HCWs have experienced an estimated 1.6-to 3.4-fold higher risk of infection relative to the general population [5,6]. Long-term care facilities (LTCFs) have been most severely impacted, with one-to two-thirds of COVID-related deaths in Europe by May 2020 estimated to have occurred among LTCF residents [7].

Healthcare facilities have undergone extensive organizational changes to combat SARS-CoV-2 transmission, particularly during local surges in COVID-19 cases. Many facilities have adopted non-pharmaceutical infection prevention and control (IPC) measures, including social distancing, reinforced contact precautions and hand hygiene procedures, use of personal protective equipment (PPE), banning of visitors, and systematic test-trace-isolate protocols. HCWs and residents of LTCFs have also been among the first populations targeted for vaccination. However, despite these interventions, nosocomial COVID-19 risk has not been eliminated, as evidenced by ongoing outbreaks in healthcare facilities worldwide. A key barrier to effective COVID-19 prevention in healthcare settings is imperfect understanding of transmission routes among patients and HCWs, and of the relative impacts of different control strategies, which depend on setting-specific organizational and demographic characteristics, immunological histories of the specific population concerned, and virological properties of locally circulating variants.

Throughout the pandemic, mathematical models (Box 1) have proven useful both to better understand the epidemiological processes underlying SARS-CoV-2 outbreaks and to support public health decision-making. Modelling studies focusing on nosocomial risk in particular, although less publicized than those focusing on community risk, have addressed a broad range of epidemiological questions [8] and aided public health decision-making [9]. However, epidemiological insights and public health recommendations from nosocomial SARS-CoV-2 models have not previously been reviewed or synthesized systematically. Here, we systematically search and review mathematical models developed to investigate SARS-CoV-2 transmission dynamics and control strategies in healthcare settings over the critical phase of the pandemic, present their main contributions, synthesize their conclusions, and discuss their limits.

## Methods

We conducted a systematic search and review of mathematical/mechanistic models of inter-individual SARS-CoV-2 transmission within healthcare settings published up to February 14, 2022. Details on the search, inclusion and exclusion criteria, screening process and data extraction are provided in the Supplementary File, and results are reported according to the PRISMA guidelines [10] (see Supplementary File).

## Results

### Model characteristics

Overall, our search identified 1,431 studies, of which 59 were included after title, abstract and full-text screening (Suppl. Fig. S1 and Suppl. Table). Most (43/59) were posted in a publicly accessible pre-print server, with a median delay of 8 months (range: 1 to 24 months) between initial preprint posting and publication.

The majority of models were stochastic (85%; 50/59), and about half were agent-based or network models (53%; 31/59). At early stages of the pandemic, when testing resources were highly limited, most studies focused on surveillance and healthcare organization (Fig. 1A). By comparison, impacts of PPE have been assessed less frequently, and vaccination strategies only began to be explored towards the end of 2020, as the first vaccines became available.

**Figure 1.**
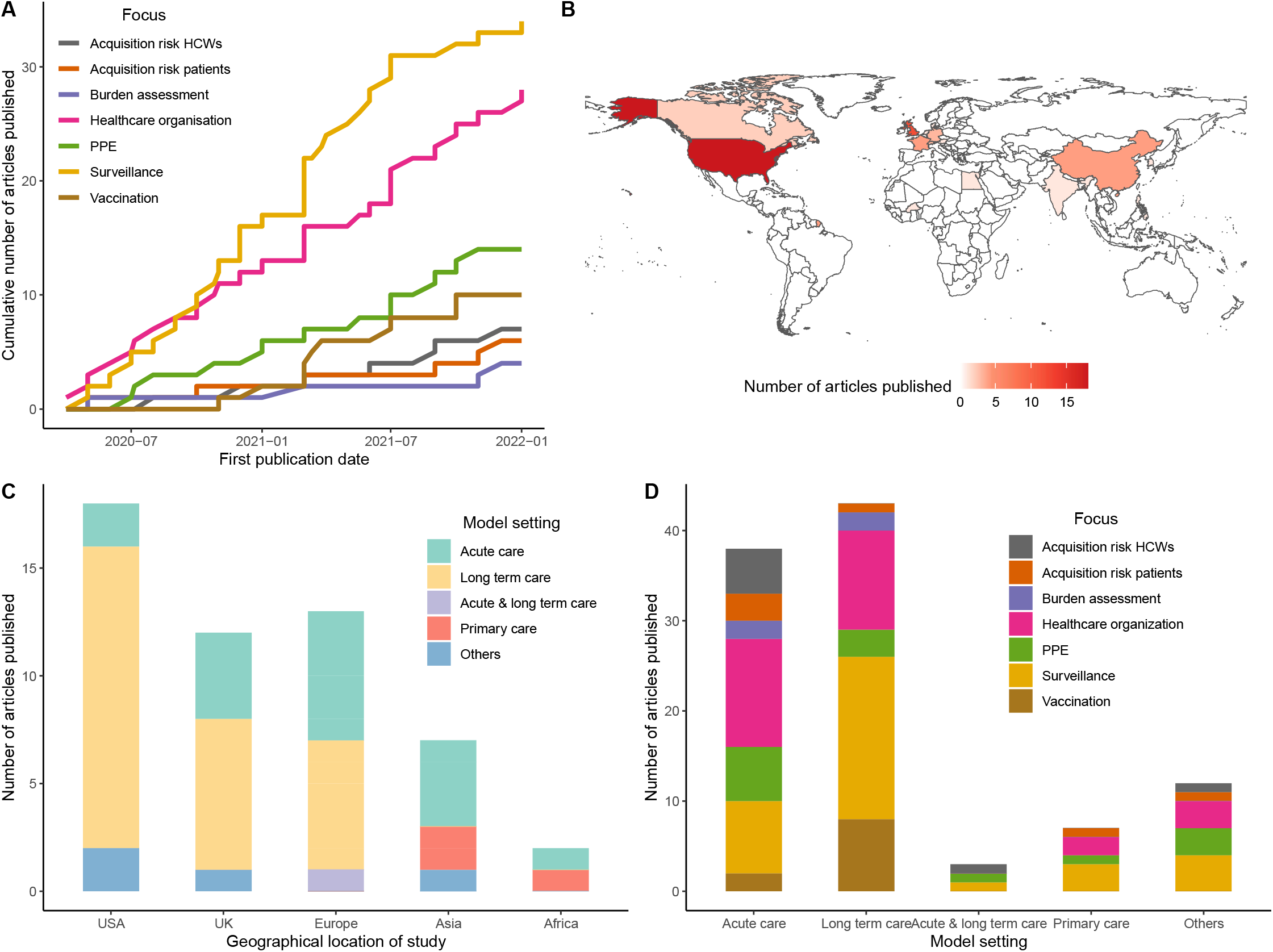
Characteristics of identified models of SARS-CoV-2 transmission within healthcare settings. (A) Cumulative number of modelling studies made accessible per month since March 2020, stratified by primary focus(es) addressed. The date used is the date of first publication, either on a public archive or in a journal. Studies addressing several subjects are counted several times. (B) Geographical distribution of countries on which the modelling is focused. (C) Distribution of modelled healthcare settings per studied country. (D) Distribution of addressed subjects, depending on the type of healthcare setting. Studies addressing several subjects are counted several times.

While models were mostly developed by teams from the USA, the UK, and other European countries, these models explored locations representing a wider range of countries worldwide (Fig. 1B). Acute care facilities and long-term care facilities were the main types of healthcare facilities considered, although this varied depending on the country of study (Fig. 1C) and the subject addressed (Fig. 1D).

### Insights on SARS-CoV-2 acquisition routes and transmission risk

Few studies have attempted to estimate the reproduction number of SARS-CoV-2 in healthcare settings, despite evidence that nosocomial and community risk may differ widely [11]. Estimates of nosocomial *R*_0_ range from 0.45 (0.36-0.56) in English acute care hospitals using a relatively simple approach [12] to 8.72 (5.14-16.32) in a French LTCF using a stochastic dynamic model accounting for imperfect surveillance [13]. Interestingly, in the latter study, *R*_0_ was estimated to drop to 1.33 (0.68-2.04) after introduction of control measures.

Several models have quantified the relative burden of SARS-CoV-2 infection and transmission among patients and HCWs over the course of the pandemic. HCWs were identified as being at high risk of occupational exposure to infection, either from colleagues or patients [14–16]. During the first wave in early 2020, they have been estimated to be the most important source of onward nosocomial transmission, both to patients and other HCWs [17,18]. However, other studies found that patient infection could result primarily from transmission from other patients [4,16,18].

### Insights on SARS-CoV-2 infection control

#### Evaluating and optimizing surveillance strategies

Models have been widely used to assess and compare the epidemiological impacts of SARS-CoV-2 testing strategies. Because SARS-CoV-2 spreads extensively through pre-symptomatic and asymptomatic transmission [19,20], the identification of non-symptomatic infections is a key component of nosocomial IPC. Several studies have highlighted difficulty controlling outbreaks when targeting only symptomatic individuals for testing [21–28]. Limited impact of only testing patients upon their admission has also been identified, suggesting that more thorough screening methods are required to limit SARS-CoV-2 introductions from the community, visitors, HCWs or ancillary staff [24,27,29,30].

Many studies have evaluated the impact of routine testing of non-symptomatic individuals. The most universal finding is that more frequent testing leads to greater reductions in nosocomial transmission [12,14,21,23,24,30–40]. Similarly, increasing daily testing capacity has been found to limit nosocomial transmission [27,41]. In the context of limited test availability early in the pandemic, effective strategies identified for optimizing nosocomial outbreak detection include pooling samples via group testing [27] and distributing tests over several batches instead of using them all at once [42].

Modelling results are less consistent concerning which subpopulations to target for routine non-symptomatic testing. Many conclude that targeting HCWs is most effective [12,25,32,43], while others suggest targeting facility patients or residents [22,27,39]. Divergence owes to underlying modelling assumptions on how patients and HCWs differ, regarding their infectiousness, susceptibility to infection, contact behaviour, and degree of interaction with visitors and other individuals in the community. For instance, testing staff proved more effective in a model of English care homes where the main source of SARS-CoV-2 introductions was staff members infected in the community [25]. Conversely, testing patients was more effective in models of a French rehabilitation hospital in which high rates of contact among ambulatory patients translated to high rates of patient-to-patient transmission [27,39]. In nursing homes, patient testing likely becomes increasingly important when visitors or other interactions with the community are permitted [33].

Lastly, in addition to testing frequency, studies have quantified the critical impact of the sensitivity and turnaround time of the test being used [24,28,30,32,33,37,38,44–47]. Several studies have identified that gains in turnaround time tend to outweigh gains in test sensitivity, explaining why rapid diagnostic tests (e.g. Ag-RDT) may be more effective than laboratory-based tests (e.g. RT-PCR) for routine non-symptomatic testing [23,25,32,39]. Conversely, if same-day test results are achievable, or if individuals effectively isolate while awaiting test results, more sensitive laboratory tests likely outperform rapid tests [33,34,44].

#### Evaluating impacts of personal protective equipment (PPE)

Several studies have found that, when available, PPE use is highly effective for reducing infection risk among both HCWs and patients. Although predicted reductions in infection risk naturally depend on assumptions underlying PPE’s impact on viral transmissibility, which vary considerably across studies and for which data are sorely lacking, several studies suggest that widespread PPE use remains a key SARS-CoV-2 prevention strategy, even when conferring comparatively low levels of protection [15,29,37,48–50]. By preventing infection, PPE use has also been shown to reduce HCW workplace absence [37] and workday loss [22]. Very few studies have tackled the question of who should be given priority to PPE access when in limited supply, although PPE has been shown to be particularly effective when accessible to all HCWs [48].

Regarding different types of PPE that may be used, Hüttel et al. [15] found hand sanitizer to be an effective means of reducing risk as a supplement to other strategies. Regarding timing, earlier introduction of PPE was found to allow a much more efficient response [22] and to enable prevention of large outbreaks [25]. However, further analyses suggest that the level of protection PPE confers can be occasionally overwhelmed in the context of large numbers of infected people in a room [51]. Finally, waning PPE adherence due to pandemic fatigue could significantly impact the efficacy of PPE-based interventions [52].

#### Evaluating and optimizing healthcare organization

Many modelling studies have assessed the epidemiological impacts of healthcare facilities adapting their structures of care and workplace organization. Larger facilities have been found to be at greater risk of nosocomial SARS-CoV-2 outbreaks [43], and splitting facilities into smaller independent units has been shown to reduce the total number of infected individuals [36,48], except when transmissibility is high [53]. Forbidding visitors was identified as not having a significant effect on outbreak probability [25], except when infection prevalence among visitors’ contacts in the community is higher than that of HCW community contacts [54].

Models have highlighted that rapid isolation of positive cases is an effective strategy for infection prevention [36,55,56]. Interestingly, models suggest that intermixing recovered individuals with potentially susceptible individuals could reduce outbreak size [32], and that sufficient spacing between patient beds is needed to limit transmission risk [36]. Results are less consistent regarding isolation upon admission. Models have highlighted the efficacy of isolating all newly admitted patients for a given duration [29] or while awaiting test results [12]. Conversely, others report no additional benefit of front-door screening or quarantine upon admission when other strategies are already in place [43,55].

Regarding staff organization, models have demonstrated benefits of cancelling HCW gatherings [57,58] and of forcing shorter and fewer patient-HCW interactions [51,58], although surprisingly this latter result was not confirmed by others [22]. Reducing between-ward staff sharing also seems to reduce both the number of wards with infected individuals [55] and the global reproduction number [37]. More complex staffing strategies have also shown potential benefits, like splitting staff into two teams that do not interact [59,60], synchronizing rather than staggering staff rotations [61], or immunity-based staffing [32,35], e.g. assigning recovered staff to infected patients [35]. Finally, admitting all COVID-19 patients to specialized quarantine hospitals in which HCWs continuously resided for 1-to-2 week-long shifts did not necessarily increase occupational HCW risk [62].

#### Evaluating vaccination strategies

All models exploring vaccination found that it could help reduce COVID-19 morbidity and mortality, especially in LTCFs [23,28,31,33,52,63–67]. However, some studies also noted that vaccination benefits could be hindered by high levels of community SARS-CoV-2 circulation [31,65] or by reduced adherence to contact precautions within facilities concomitant with vaccine rollout, for instance due to pandemic fatigue or risk compensation [52].

A major focus of these models has been evaluation of which groups of individuals to target or prioritize for vaccination in a context of limited vaccine availability, yielding sometimes inconsistent results. Some found that LTCF residents should be prioritized over staff, especially in LTCFs with low adherence to IPC measures [33,52]. Conversely, staff vaccination was shown to be particularly effective for decreasing the overall attack rate, especially in the absence of a testing protocol [64]. Targeting staff for vaccination may also be preferable when the risk of virus importation from the community is high [31]. Finally, it has been shown that vaccine rollout should first target staff members most exposed to potential COVID-19 patients (e.g. staff from emergency departments) [63].

It should be noted that the conclusions reached by these models depend strongly on modelling assumptions underlying vaccine action. Across models, assumed mechanisms related to vaccination effectiveness included one or several of the following: a reduction in acquisition risk, a reduction in symptom risk, and a reduction in the infectiousness of infected vaccinated individuals. For instance, it was shown that if a vaccine only reduces symptom risk, then increasing vaccination among nursing home staff leads to an increase in the proportion of asymptomatic infections among staff, which subsequently leads to increased infection risk for residents [23]. Additionally, no model considered vaccine impact over the long-term, which is particularly relevant in the context of waning immunity and the emergence of novel variants with vaccine-escape properties.

## Discussion

Mathematical models have become ubiquitous tools to help understand the dynamics of infectious disease outbreaks and to support public health decision-making. Here, we have reviewed how models have helped to inform COVID-19 risk management in healthcare settings, in particular by providing a better understanding of nosocomial transmission dynamics and control strategy effectiveness.

The generation of *in silico* evidence to inform infection control strategies has been the leading motivation for nosocomial SARS-CoV-2 transmission modelling. Although real-world evidence collected during randomized controlled trials is the gold-standard, such data are extremely difficult to generate in the context of a rapidly evolving epidemic. Beyond the obvious costs and time involved, great heterogeneity in population characteristics and exposure risk across different healthcare settings means that a large number of centres must be enrolled to achieve sufficient cluster randomisation. Several high-impact randomized controlled trials have nonetheless been successfully conducted in healthcare settings despite these challenges, in particular to evaluate COVID-19 vaccines, therapies and pre- or post-exposure prophylactic agents [68–71]. However, trials evaluating impacts of common IPC interventions, including surveillance testing, PPE and healthcare reorganization, are scarce [72,73].

In this context, mathematical modelling approaches have been particularly helpful to investigate critical IPC questions in (near) real-time, since they allow for the simulation of diverse scenarios at relatively high speed and low cost, while accounting for all available knowledge and uncertainty at a given place and time. Model-based evaluations also allow for levels of granularity in intervention arms that may be infeasible in real trial designs. Our review highlights the range of modelling studies published before the end of 2020, at a time when the scientific and medical communities were in particularly great need of evidence to inform optimal allocation of limited infection prevention resources, including RT-PCR tests, face masks and, later, vaccines.

However, two common means of SARS-CoV-2 transmission prevention with important implications for the field have been notably under-addressed. First, modelling studies on the relative impact of different types of face masks (e.g. surgical masks, N95 respirators) are scarce [74], tied to a lack of precise data on how they impact viral transmissibility, as well as their potential indirect roles as transmission vectors. Second, although ubiquitous in practice at various stages of the pandemic, explicit social distancing interventions have rarely been assessed [22,39]. This is probably because accurate modelling of social distancing requires fine-scaled simulation of inter-individual contact networks, typically using an agent-based approach, which is beyond the scope of most models. When faced with both epistemic uncertainty and a need for relative computational simplicity, modelers may be reluctant to include and formalize specific interventions that require arbitrary, highly sophisticated and/or potentially wrongheaded assumptions about their mechanistic impacts on SARS-CoV-2 transmission. Instead, a common modelling strategy has been to include generic non-pharmaceutical interventions that simply reduce the viral transmission rate, and which are assumed to represent any combination of basic infection prevention interventions, including face masks, gloves, gowns, face shields, hand hygiene or social distancing.

Relative to the evaluation of infection control strategies, modelling studies have more rarely focused on the estimation of epidemiological parameters using statistical inference. In particular, in the event of the sudden emergence of a novel infectious disease, *R*_0_ estimation is essential for epidemic forecasting and emergency response planning, and relies largely on mathematical modelling approaches. Although estimates of *R*_0_ quickly became available for SARS-CoV-2 across various community settings in early 2020, [75], estimates specific to healthcare settings remain scarce. Yet there is a great need for robust estimates across diverse settings, as underlying levels of epidemic risk vary substantially across facilities due to their intrinsic heterogeneity (e.g. types of care provided, population sizes, contact behaviour of these populations). For instance, assuming *R*_0_=3.5 in the community, Temime et al. [11] estimated that nosocomial *R*_0_ could range from 0.7 to 11.7, depending on the type of ward and the density of contacts among and between patients and HCWs. This heterogeneity in nosocomial *R*_0_ is consistent with the range of estimates derived from models described in this review [12,13], and has critical implications, informing which facilities and populations are most at risk for explosive outbreaks and thus most in need of urgent infection control measures.

This lack of evidence likely stems from both data limitations and remaining uncertainty about the relative importance of precise paths of transmission (e.g. through direct person-to-person contact; transient viral carriage on hands, garments or shared medical devices; stagnant air in poorly ventilated spaces). Particularly early in the pandemic, nosocomial COVID-19 data came primarily from contexts of emergency outbreak management rather than routine data collection through stable surveillance systems. For future waves of SARS-CoV-2 and other infectious diseases, the estimation of epidemiological parameters may be made easier by harnessing large databases that systematically gather patient and HCW tests results, administrative data and healthcare exposures across diverse healthcare facilities over time.

Researchers have faced significant challenges when developing SARS-CoV-2 transmission models. First, data limitations, particularly early in the pandemic, forced many modelers to make assumptions that oversimplify healthcare facility structure, population behaviour and SARS-CoV-2 transmission dynamics, limiting the applicability of some results to real-world settings. Second, the shifting epidemiological landscape – characterized not only by the rapid spread of SARS-CoV-2, but also rapid change in population behaviours, sudden resource shortages, consecutive changes in public health recommendations, rapid approval of novel diagnostics, therapies and vaccines, and the successive emergence of distinct variants of concern – required researchers to continually adapt their models in order to remain useful, with relevant data required to parameterize these updates often lagging behind.

Greater interdisciplinarity will be required to maximize the utility of mathematical modelling in the future. More direct lines of contact between modelers, hospital infection control teams, clinicians and decision-makers should guide modelers in their research. First, this may help to steer studies towards the questions that are most clinically relevant, as informed by the needs and issues faced in real clinical settings. Second, this may help modelers to evaluate strategies that are feasible in practice, considering logistical constraints such as workforce or equipment availability and hospital structure. Third, these discussions may inspire modelers to account for outcomes beyond transmission risk and infection burden, such as cost-effectiveness or mental health. Indeed, interventions such as visitor restrictions or staff re-organization can have a great impact on the social isolation of patients or workload of HCWs, which is difficult to take into account explicitly in mathematical models. Cost-effectiveness is increasingly considered in modelling studies; for instance, several studies have quantified the health-economic efficiency of frequent non-symptomatic testing [12,24,38,39,45]. However, more frequent estimation of health-economic outcomes may increase their usefulness for decision-makers, who must balance the competing priorities of maximizing population health and minimizing monetary cost. Finally, increased communication across disciplines may facilitate more timely sharing of modelling results to those who may benefit from them most, including infection control teams and hospital administrators. The use of social media and the surge in posting of articles on pre-print repositories during the COVID-19 pandemic have helped to facilitate the timely sharing of results, but there remains an onus on academic publishers to ensure a timely peer review process so that modelling results are shared quickly enough to maximize their impact.

This review has several limitations. First, we chose to exclude all statistical, mathematical, or computational models not including inter-individual SARS-CoV-2 transmission. Consequently, other types of models such as physical or biomechanical models of airborne transmission were excluded [76–78]. Second, we excluded articles posted on public archives such as arXiv, medrXiv or biorXiv [79], which are not subject to peer review and can be difficult to track. Although we did include some preprints in our review, we were unable to systematically explore these archives. Third, we may have missed articles published in journals not referenced in PubMed (e.g. computer science or mathematical journals). However, since our focus was on epidemiological insights and public health recommendations, we do not believe that this significantly impacted our main findings.

## Conclusion

Often developed in the face of great epidemiological uncertainty, mathematical models have come to form a rich evidence base describing how SARS-CoV-2 spreads in healthcare settings and informing which nosocomial COVID-19 control strategies are optimal, in particular with regards to healthcare reorganization and the allocation of limited supplies of PPE, diagnostic testing and vaccines. Into the future, epidemiological models may continue to inform control strategies for outbreaks of SARS-CoV-2 and other viral respiratory pathogens, but increased collaboration should be sought between modelers, hospital infection control teams, clinicians, and public health decision-makers to help maximize their utility.

## Data Availability

All data produced in the present work are contained in the manuscript

## Conflict of interest

Authors report no conflict of interest.

## Funding

This study received funding from the MODCOV project from the Fondation de France (Grant 106059) as part of the alliance framework “Tous unis contre le virus”, the Université Paris-Saclay (AAP Covid-19 2020) and the French government through its National Research Agency project SPHINX-17-CE36-0008-01 and the “Investissement d’Avenir” program, Laboratoire d’Excellence “Integrative Biology of Emerging Infectious Diseases” (Grant ANR-10-LABX-62-IBEID). The work was also supported directly by internal resources from the French National Institute for Health and Medical Research (Inserm), Institut Pasteur, le Conservatoire National des Arts et Métiers, and l’Université Versailles Saint-Quentin-en-Yvelines/Université Paris-Saclay.

## Author contributions

LO, LT, SK conceived the study, LO and LT acquired the funding. Screening of the articles was performed by DRMS, SC, GS, SJ, KJ, LO and LT. Data extraction and analysis were performed by DRMS, SC, GS, SJ, LO and LT. DRMS, SC and TP rendered figures and statistics. All authors contributed to writing.

## Figure captions

***Box 1: Mathematical models of SARS-CoV-2 transmission in health care settings***

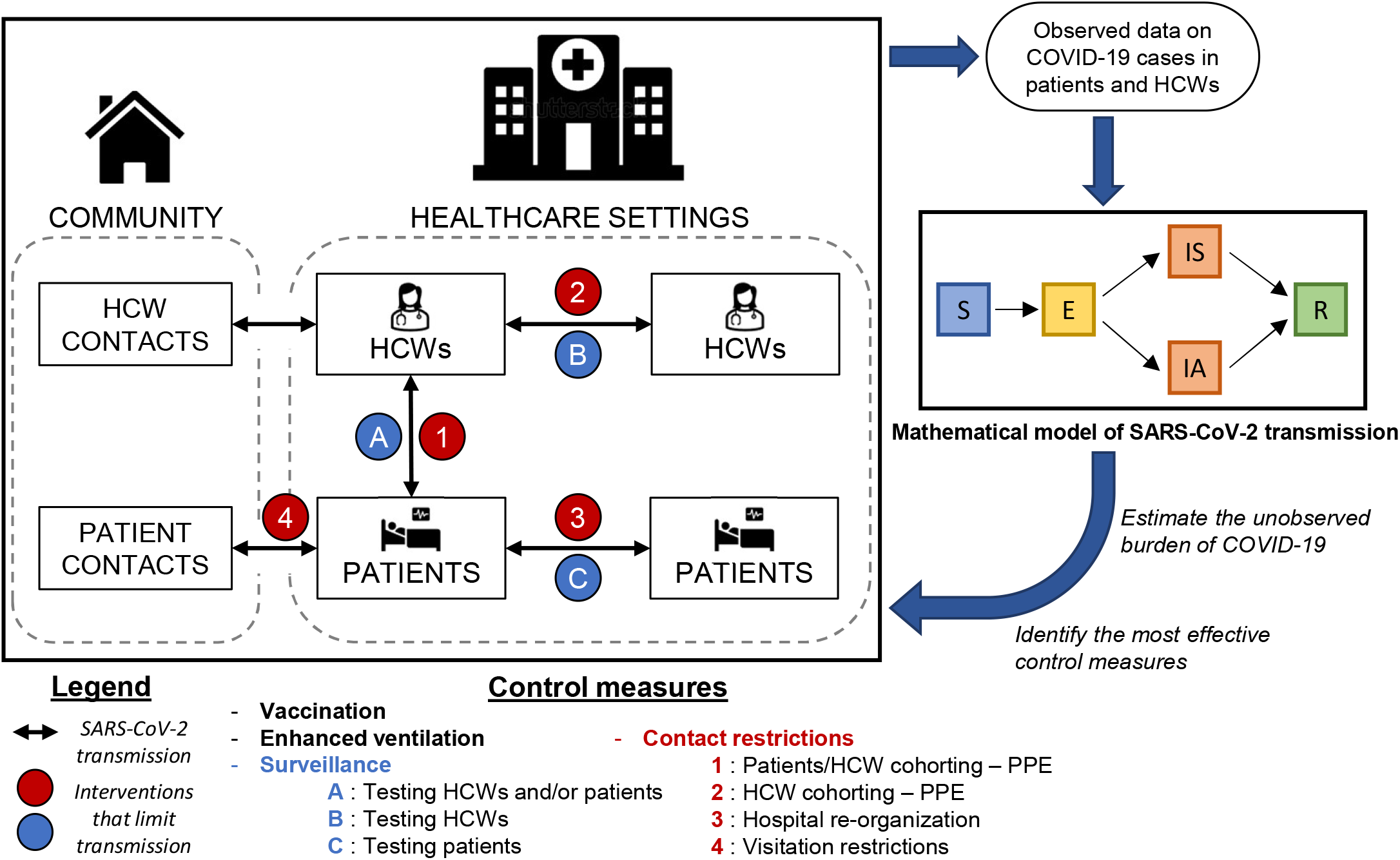

Mathematical models are theoretical constructs that mechanistically formalize the dynamic processes underlying SARS-CoV-2 transmission. A typical model splits the individuals present in a healthcare facility into sub-populations, including one or more categories of patients (or residents) and HCWs. Mathematical or statistical tools are used to describe the natural history of infection through the definition of different infection states or “compartments”. For instance, in the specific case of SARS-CoV-2, which is characterized by an incubation period subsequent to exposure and the acquisition of (partial) immunity after infection, the main compartments considered are: Susceptible to infection (S), Exposed to infection or incubating (E), Infectious (I), and Recovered or immunized (R). As a large share of infectious individuals may be asymptomatic, the I compartment is often subdivided into asymptomatic (IA) and symptomatic (IS) compartments. Various other sources of heterogeneity may also be considered, including different levels of viral shedding among infectious individuals, or different trajectories of care among symptomatic individuals (e.g., isolation, mechanical ventilation, admission to intensive care, death). Specific contact patterns between individuals of different sub-populations and infection statuses can further be accounted for through the definition of contact matrices.

Compartmental models are most frequent, but agent-based models (also known as individual-based models) are another common formalism, in which each unique individual in the population is explicitly modelled. This enables more detailed integration of heterogeneity in contact patterns, disease progression, transmission risk and other epidemiological processes. Models can further be categorized as either deterministic or stochastic. In deterministic models, there is no randomness in epidemiological processes, and a particular set of initial conditions always results in identical outbreaks. By contrast, in stochastic models, it is possible to account for randomness in the parameters or processes included, resulting in different outbreak trajectories each time the model is run. Stochasticity is particularly relevant in models of healthcare settings, where population sizes are small and randomness can have a strong impact on outbreak dynamics.

Models are used for a variety of purposes. They are widely used to simulate virus transmission in specific settings and populations, allowing for the quantification of virus burden in particular epidemiological scenarios (e.g. after the introduction of a novel SARS-CoV-2 variant into a hospital via newly admitted patients, short-stay visitors, or members of staff infected in the community). Models are also used to enable the *in silico* assessment of public health interventions through the mechanistic implementation of interventions (e.g. testing, isolation, PPE provisioning, contact restrictions, vaccine deployment). Intervention impact can then be evaluated by simulating counterfactual outbreaks with and without the intervention in place. Finally, models are key to analysing reported data from real outbreaks (e.g. time series data, individual line lists of SARS-CoV-2 cases), accounting not only for unobserved processes (e.g., virus transmission) but also incomplete infection data, whether due to the presence of asymptomatic infections, imperfect reporting, or limited surveillance capacity. This allows for the retrospective assessment of true disease burden from a given outbreak (e.g., cumulative infection incidence) as well as estimation of important epidemiological parameters such as *R*_0_, the basic reproduction number, which describes the average number of secondary cases caused by an index case in an immunologically naïve population. *R*_0_ is particularly helpful to understand the epidemic potential of an emerging pathogen, though its value may vary across distinct sub-populations and settings, such as particular groups of patients and HCWs in particular healthcare facilities.

## Notes

### Competing Interest Statement

The authors have declared no competing interest.

### Summary of Updates

Figures updated with legends

