## Supplementary File for "How have mathematical models contributed to understanding the transmission and control of SARS-CoV-2 in healthcare settings? A systematic search and review"

**Summary of contents:**

- **Methods**
- **Figure S1:** Flowchart diagram of study selection using PRISMA flow Diagram recommendations
- **Table:** Data extracted for all articles included
- **PRISMA Checklist**

**METHODS**

***Search query***

We searched PubMed for original research articles published until February, 14, 2022 using the following search query:

("outbreak*"[Title/Abstract] OR "dynamic*"[Title/Abstract] OR

"disseminat*"[Title/Abstract] OR "propagat*"[Title/Abstract] OR

"transmi*"[Title/Abstract] OR "cross infection/transmission"[MeSH Terms] OR

"spread*"[Title/Abstract] OR "diffus*"[Title/Abstract] OR

"circulati*"[Title/Abstract] OR "epidemiology"[Title/Abstract])

AND

("model*"[Title/Abstract] OR "models, theoretical"[MeSH Terms] OR "simulat*"[Title/Abstract])

AND

("mathematic*"[MeSH Terms] OR "compartmental"[Title/Abstract] OR

"agent-based"[Title/Abstract] OR "individual-based"[Title/Abstract] OR

"stochastic*"[Title/Abstract] OR "deterministic"[Title/Abstract] OR

"computer-generated"[Title/Abstract] OR "computer simulation"[MeSH Terms] OR

"computer"[Title/Abstract] OR "computational"[Title/Abstract] OR

"mathematical*"[Title/Abstract] OR "theoretical"[Title/Abstract] OR

"prediction"[Title/Abstract] OR "simulat*"[Title/Abstract])

AND

("hospital*"[Title/Abstract] OR "ward*"[Title/Abstract] OR

"healthcare"[Title/Abstract] OR "health care"[Title/Abstract] OR

"facility"[Title/Abstract] OR "facilities"[Title/Abstract] OR

"long-term care"[Title/Abstract] OR "retirement home"[Title/Abstract] OR

"nursing homes"[Title/Abstract] OR "nursing home"[Title/Abstract] OR

"nosocomial"[Title/Abstract])

AND

("COVID-19"[Title/Abstract] OR "SARS-CoV-2"[Title/Abstract])

***Inclusion and exclusion criteria***

Studies were eligible if they: (i) focused on SARS-CoV-2 spread within healthcare settings (hospitals; hospital units, wards, or bays; nursing/retirement homes or long-term care facilities; primary/community healthcare centres or testing centres); and (ii) included a dynamic mathematical/mechanistic inter-individual transmission model (compartmental, individual-based or network model).

Studies were excluded if they: (i) only presented statistical models, epidemiologic data, or models not involving inter-individual SARS-CoV-2 transmission; (ii) focused on aerial modelling of fluid mechanics; or (iii) did not present original results.

***Article screening***

Article selection was conducted in two steps. First, each article retrieved from the search query was independently screened using the above selection criteria by two reviewers on the basis of its title and abstract. Second, screened articles were subject to full-text assessment to confirm their eligibility. Screening and full-text assessment were conducted using the Covidence review tool [1], whereby articles were randomly distributed among contributing authors for both screening and full-text assessment. Any conflicts in article screening or full-text assessment were resolved through discussion of the entire group of authors.

***Data extraction and analysis***

We conducted a descriptive analysis of the identified articles. Data extracted for each article by one author included: date of publication; type of model (e.g. deterministic compartmental, stochastic agent-based); main research objectives; country of study when applicable; type of healthcare facility (e.g. hospital, LTCF); and population studied (e.g. patients, HCWs). The main findings from each article were then extracted and summarized, focusing on three questions: (i) how did the study contribute to understanding SARS-CoV-2 transmission and acquisition risk within healthcare settings? (ii) which (if any) public health interventions were evaluated by the model? and (iii) what did the study conclude regarding the epidemiological impact of evaluated interventions?

[1]    Covidence systematic review software. Veritas Health Innovation, Melbourne, Australia n.d. https://www.covidence.org/ (accessed January 20, 2023).

***Suppl Fig S1: Flow chart diagram of study selection using PRISMA Flow Diagram recommendations***

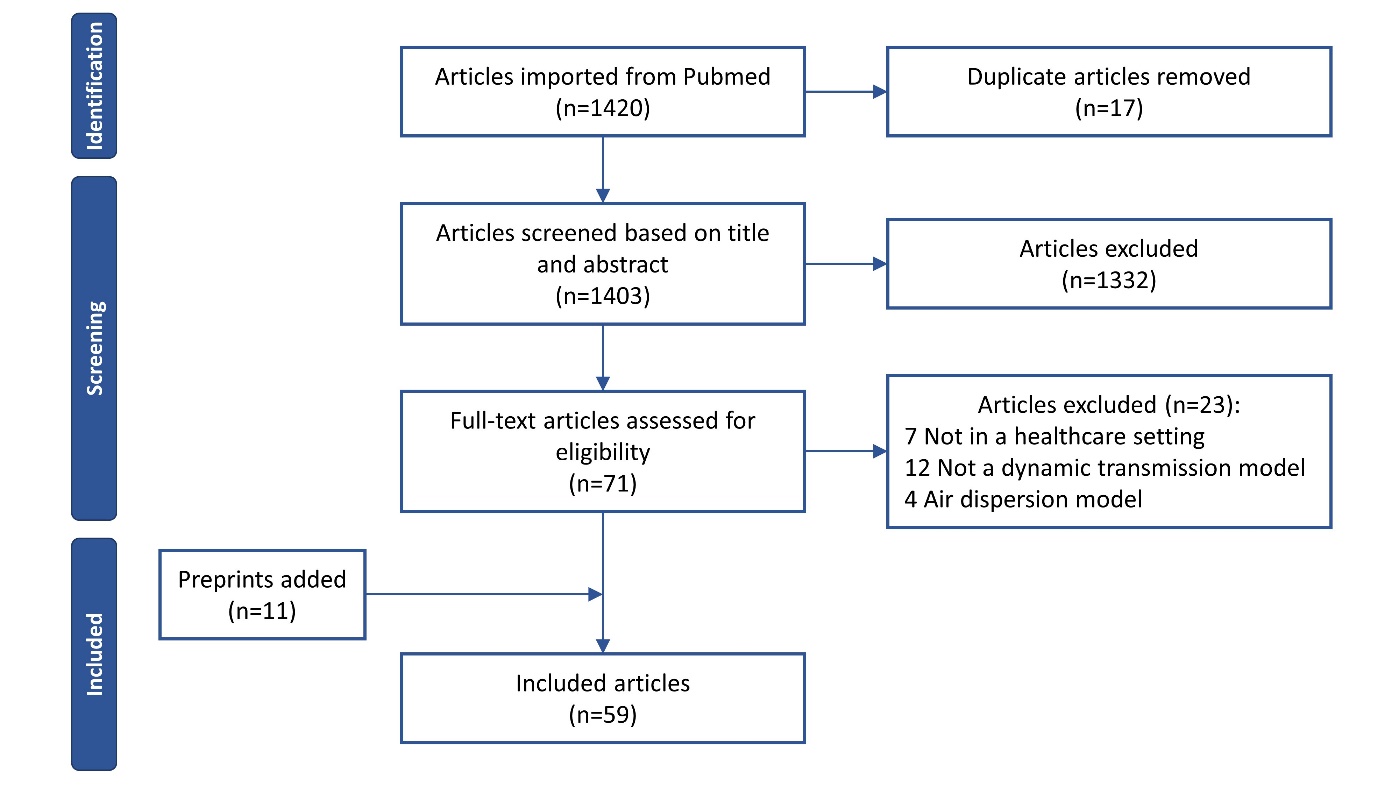

***Suppl Table: Complete list of identified dynamic models of SARS-CoV-2 transmission within healthcare settings***

| **First author** | **Title** | **Publi Date** | **Journal** | **Type of model** | **Main objectives** | **Country of study** | **Type of facility** | **Population of interest** |
| --- | --- | --- | --- | --- | --- | --- | --- | --- |
| Abbas | Explosive nosocomial outbreak of SARS-CoV-2 in a rehabilitation clinic: the limits of genomics for outbreak reconstruction | Nov 2021 | Journal of Hospital infection | Stochastic Compartmental | Reconstruct transmission chains over a nosocomial outbreak | Switzerland | Rehabilitation clinic | Patients and HCWs |
| Baek | A mathematical model of COVID-19 transmission in a tertiary hospital and assessment of the effects of different intervention strategies | Oct 2020 | PLoS ONE | Deterministic Compartmental | Evaluate screening and quarantine upon admission, early testing of suspected cases, and personal protective equipment for staff and visitors | South Korea | Tertiary hospital | Patients and HCWs |
| Blumberg | Modeling scenarios for mitigating outbreaks in congregate settings | July 2022 | Plos Computational Biology | Stochastic Agent-based | Explore how interventions that decrease the size of the susceptible populations, such as vaccination or depopulation, impact the expected number of infections | California | Congregate settings, such as prisons or nursing homes. | Residents |
| Bosbach | Maximization of Open Hospital Capacity under Shortage of SARS-CoV-2 Vaccines—An Open Access, Stochastic Simulation Tool | May 2021 | Vaccines | Stochastic Agent-based | Explore the effect of different staff vaccination rollout schemes | Germany | Hospital | HCWs |
| Cardinal | Simulating a Community Mental Health Service During the COVID-19 Pandemic: Effects of Clinician-Clinician Encounters, Clinician-Patient-Family Encounters, Symptom-Triggered Protective Behaviour, and Household Clustering. | Feb 2021 | Frontiers in psychiatry | Stochastic Agent-based | Examine effects of type of appointment and clinician-clinician encounters on infection rates | / | Mental health facility | Patients and HCWs |
| Chen | Bayesian inference of heterogeneous epidemic models: Application to COVID-19 spread accounting for long-term care facilities. | Nov 2021 | Computer Methods in Applied Mechanics and Engineering | Deterministic Compartmental | Fit a model incorporating many dimensions including heterogeneity using Bayesian methods | USA, New Jersey and Texas | Within and outside LTCF | Entire population |
| Cheng | Measures to prevent nosocomial transmissions of COVID-19 based on interpersonal contact data | Jan 2022 | Primary Health Care Research and Development | Stochastic Compartmental | Assess the impact of a medical staff rotation system, the establishment of a separate fever clinic and medical staff working alone | Hebei province (China) | Hospital | Patients and HCWs |
| Chin | Frequency of routine testing for Coronavirus Disease 2019 COVID-19 in high-risk healthcare environments to reduce outbreaks | Nov 2021 | CID | Stochastic Compartmental | Assess the impact of various testing frequencies in a healthcare setting population | / | High risk healthcare environment (LTCF, hospital) | Patients and HCWs with no distinction |
| Cooper | The burden and dynamics of hospital-acquired SARS-CoV-2 in England | Nov 2021 | Research Square | Deterministic Compartmental | Quantify within-hospital transmission, evaluate likely pathways of spread and explore the dynamical consequences in the community. | UK | Hospitals | Patients and HCWs (+ general community) |
| Ding | Surveillance Testing for Rapid Detection of Outbreaks in Facilities | Oct 2021 | ArXiv | Stochastic Agent-based | Identify the best testing strategies to detect SARS-CoV-2 in facilities such as nursing homes | USA | Nursing homes or meat-packing plants | Residents and staff |
| Dy | A COVID-19 infection risk model for frontline health care workers | Aug 2020 | Netw Model Anal Health Inform Bioinform | Deterministic Compartmental | Examine the risk factors of virus transmission per day, quantify these risks, estimate the number of new infections, and suggest ways to minimize these risks | Philippines | Hospital | HCWs |
| Evans | Quantifying the contribution of pathways of nosocomial acquisition of COVID-19 in English hospitals | Dec 2021 | Int J Epidemiol | Stochastic Agent-based | Quantify relative contribution of routes of transmission (between community, HCWs and patients, directly and indirectly) to nosocomial infections | UK | Hospitals | Patients and HCWs |
| Evans | The impact of testing and infection prevention and control strategies on within- hospital transmission dynamics of COVID-19 in English hospitals | May 2021 | Philosophical Transactions of the Royal Society B | Deterministic Compartmental | Assess the impact of periodic staff testing and patient case isolation | UK | Hospital | Patients and HCWs |
| Ferris | Efficacy of FFP3 respirators for prevention of SARS-CoV-2 infection in healthcare workers. | Nov 2021 | Elife | Stochastic other | Quantify effectiveness of switching from fluid-resistant surgical masks to FFP3 and check increased infection risk for HCWs working in covid wards | UK (NHS) | Tertiary hospital | HCWs |
| Fosdick | Model-based Evaluation of Continued COVID-19 Risk at Long Term Care Facilities | Sept 2022 | Infectious Disease Modelling | Stochastic Agent-based | Quantify the continued risk for COVID-19 infections within a facility given a designated testing schedule and vaccine requirement | USA | LTCF | Residents and staff |
| Gomez-Vazquez | Testing and Vaccination to Reduce the Impact of COVID-19 in Nursing Homes: An Agent-Based Approach | May 2022 | BMC infectious Diseases | Stochastic Agent-based | Quantify the effect of testing and vaccine strategies on the attack rate, length of the epidemic, and hospitalization | USA | Nursing homes | Residents and staff |
| Hall | Outbreaks in care homes may lead to substantial disease burden if not mitigated | May 2021 | Philosophical Transactions of the Royal Society B: Biological Sciences | Stochastic Compartmental | Forecast eventual potential burden in UK care homes | UK | Care homes including nursing homes | Residents |
| Hollinghurst | Intensity of COVID-19 in care homes following Hospital Discharge in the early stages of the UK epidemic | Mar 2022 | Age Ageing | Stochastic other | Assess the impact of hospital discharge events in the intensity of care home cases | Wales, UK | Care homes | Residents |
| Holmdahl | Modeling the impact of vaccination strategies for nursing homes in the context of increased SARS-CoV-2 community transmission and variants | Jan 2022 | Clin Inf Diseases | Stochastic Agent-based | Assess the impact of community prevalence, the Delta variant, staff vaccination coverage, and boosters for residents on outbreak dynamics in nursing homes | USA (Massachussetts) | LTCF, nursing homes | Residents and staff |
| Holmdahl | Estimation of Transmission of COVID-19 in Simulated Nursing Homes With Frequent Testing and Immunity-Based Staffing | May 2021 | JAMA Network open | Stochastic Agent-based | Assess the impact of contact-targeted interventions and testing | USA (Massachussetts) | LTCF, nursing homes | Residents and staff |
| Huang | SARS-CoV-2 transmission and control in a hospital setting: an individual-based modelling study | Mar 2021 | Royal Society Open Science | Stochastic Agent-based | Assess the impact of social distancing, self-isolation, tracing and quarantine, and wearing facial masks or personal protective equipment. | China | Hospital | HCWs |
| Hüttel | Analysis of social interactions and risk factors relevant to the spread of infectious diseases at hospitals and nursing homes | Sept 2021 | PLOS ONE | Stochastic Network | Identify infection risk factors among HCWs | Denmark | Hospital and nursing homes | HCWs |
| Jang | COVID-19 modeling and non-pharmaceutical interventions in an outpatient dialysis unit | July 2021 | Plos Computational Biology | Stochastic Agent-based | Show that there is a combination of simple and inexpensive NPIs that can substantially lower the impact of COVID-19 in the dialysis unit. | Iowa | Dialysis units | Patients and HCWs |
| Jijόn | Risk of incident SARS-CoV-2 infection among healthcare workers in Egyptian quarantine hospitals | Nov 2022 | Scientific reports | Stochastic Compartmental | Assess the occupational infection risk for HCWs in Egyptian quarantine hospitals | Egypt | Hospital | HCWs |
| Kahn | Mathematical modeling to inform vaccination strategies and testing approaches for COVID-19 in nursing homes | June 2021 | Clin Inf Diseases | Stochastic Agent-based | Understand the expected impact of vaccination and of different screening testing strategies in nursing homes | USA | Nursing home | HCWs |
| Kendall | Antigen-based Rapid Diagnostic Testing or Alternatives for Diagnosis of Symptomatic COVID-19: A Simulation-based Net Benefit Analysis. | Nov 2021 | Epidemiology | Stochastic other | Compare the performance of PCR vs. antigen testing for transmission prevention | / | Hospital setting with high prevalence (40%) | Not specified |
| Kluger | Impact of healthcare worker shift scheduling on workforce preservation during the COVID-19 pandemic | July 2020 | Infection Control & Hospital Epidemiology / Cambridge | Stochastic Agent-based | Model various inpatient rotation schedules of physicians and nurses to determine patterns associated with optimal workforce preservation and lower nosocomial infections in settings in which PPE is imperfect or unavailable | USA | Hospital | Patients and HCWs |
| Lasser | Agent-based simulations for protecting nursing homes with prevention and vaccination strategies | Dec 2021 | Journal of the Royal Society Interface | Stochastic Agent-based | Identify optimal mitigation testing and vaccination strategies for nursing homes | Austria | Nursing home | Residents and staff |
| Litwin | Preventing COVID-19 Outbreaks Through Surveillance Testing in Healthcare Facilities - A Modelling Study | Jan 2022 | BMC infectious Diseases | Stochastic Agent-based | Examine different surveillance testing strategies for their potential to decrease the probability of outbreaks | Germany | Hospital | Patients and HCWs |
| Liu | Susceptible-Infected-Removed Mathematical Model under Deep Learning in Hospital Infection Control of Novel Coronavirus Pneumonia. | Oct 2021 | Journal of Healthcare Engineering | Stochastic Compartmental | Understand spread within a hospital, simulate the impact of a potential vaccine and other control measures | China | Hospital | Patients |
| Love | Continued need for non-pharmaceutical interventions after COVID-19 vaccination in long-term-care facilities | Sept 2021 | Scientific Reports | Stochastic Agent-based | Evaluate the interacting effects of non-pharmaceutical interventions and vaccination | USA | LTCF | Residents and staff |
| Love | Comparison of antigen- and RT-PCR-based testing strategies for detection of Sars-Cov-2 in two high-exposure settings | Sept 2021 | PLoS ONE | Stochastic Agent-based | Investigate the population-level epidemiological effects and compare the cost-effectiveness of different testing strategies, including RT-PCR, antigen, and two reflex testing strategies | USA | Nursing home + university dormitory | Residents and staff |
| Lucia-Sanz | Modeling shield immunity to reduce COVID-19 transmission in long-term care facilities | Jan 2023 | Annals of epidemiology | Stochastic network | Study the effectiveness of shield immunity in reducing outbreak size in LTCFs | USA | LTCF | Residents and staff |
| Martos | Modelling the transmission of infectious diseases inside hospital bays: implications for Covid-19 | Nov 2020 | Math Biosci and Eng | Stochastic Agent-based | Assess the impact of bay size on COVID spread and the efficacy of case isolation and periodic testing | UK | Hospital | Patients |
| Nguyen | Evaluating intervention strategies in controlling COVID-19 spread in care homes: An agent-based model | Sept 2021 | Infection Control & Hospital Epidemiology | Stochastic Agent-based | Find control interventions that are both effective to contain spread and feasible in a care home | Scotland | Care homes | Residents and staff |
| Nguyen | Impact of visitation and cohorting policies to shield residents from covid-19 spread in care homes: an agent-based model | Sept 2021 | AJIC: American Journal of Infection Control | Stochastic Agent-based | Examine the impact of visitation, cohorting policies and care home population size upon the spread of COVID-19 and the risk of outbreak occurrence in this setting | Scotland | Care homes | Residents |
| Nguyen | Hybrid simulation modelling of networks of heterogeneous care homes and the inter-facility spread of Covid-19 by sharing staff | Jan 2022 | Plos Computational Biology | Stochastic Agent-based | Explore Covid-19 spread across multiple care homes via bank/agency staff and evaluate the effectiveness of interventions targeting this group | Scotland (UK) | Nursing homes | Residents and staff |
| Oodally | Hospital-level work organization drives the spread of SARS-CoV-2 within hospitals: insights from a multi-ward model | Sept 2021 | Medrxiv | Stochastic Compartmental | Understand the risk of SARS-CoV-2 spread in a multi-ward hospital with staff sharing, and assess control interventions | France | Psychiatric hospital | Patients and HCWs |
| Pham | Interventions to control nosocomial transmission of SARS-CoV-2: a modelling study | Aug 2021 | BMC medicine | Stochastic Agent-based | Evaluate the efficacy of PPE, screening, contract tracing for outbreaks of variants with higher transmissibility | Netherlands | Hospital | Patients and HCWs |
| Procter | SARS-CoV-2 infection risk during delivery of childhood vaccination campaigns: a modelling study. | Aug 2021 | BMC medicine | Stochastic Compartmental | Understand additional infection risk to children receiving vaccination, their caregivers and vaccinators during an outbreak. | Burkina Faso, Ethiopia, Brasil | Children vaccination center | Vaccinators, vaccinees and their caregivers |
| Qiu | Evaluating the contributions of strategies to prevent SARS-CoV-2 transmission in the healthcare setting: a modelling study | Mar 2021 | BMJ Open | Stochastic Compartmental | Assess the impact of testing, PPE and cohorting | USA | Hospital | Patients and HCWs |
| Rosello | Impact of non-pharmaceutical interventions on SARS-CoV-2 outbreaks in English care homes: a modelling study | Apr 2022 | BMC infectious Diseases | Stochastic Compartmental | During second wave with interventions already widely in place, study how probability of outbreak varies with: - different routes of introduction (staff, visitors, residents returning from hospital) - community prevalence - different testing and IPC interventions | England | Nursing homes and care homes | Residents and staff |
| Sanchez-Taltavull | Regular testing of asymptomatic healthcare workers identifies cost-efficient SARS-CoV-2 preventive measures | Nov 2021 | PLoS ONE | Deterministic Compartmental | Study how regular testing and a desynchronisation protocol are effective in preventing transmission of COVID-19 infection at work, and compare both strategies in terms of workforce availability and cost-effectiveness | Switzerland | Hospital | HCWs |
| Sánchez-Taltavull | Modelling strategies to organize healthcare workforce during pandemics: Application to COVID-19 | Aug 2021 | Journal of Theoretical Biology | Stochastic Compartmental | Identify the best organizational strategies (staff scheduling) to minimize the risk of Covid contamination for the staff | / | Hospital | HCWs |
| Saurabh | Dynamics of SARS-CoV-2 Transmission Among Indian Nationals Evacuated From Iran. | Oct 2020 | Disaster Medicine and Public Health | Deterministic Compartmental | Examine epidemic spread in a quaranting facility where individuals are tested at entry and exit | India, Jaisalmer, Rajasthan | Quarantine facility for Indians returning from abroad | Patients |
| Schmidt | Using Non-Pharmaceutical Interventions and High Isolation of Asymptomatic Carriers to Contain the Spread of SARS-CoV-2 in Nursing Homes | Jan 2022 | Life | Stochastic Compartmental | Compare the effect of R0, common NPIs, and isolation rates of pre-symptomatic carriers on SARS-Cov2 attack rate, peak cases, and timing in a 200-resident nursing home. | USA | Nursing home | Residents, staff, visitors and community members |
| See | Modeling effectiveness of testing strategies to prevent COVID-19 in nursing homes —United States, 2020 | Aug 2021 | Clin Inf Diseases | Stochastic other | Impact of serial testing (weekly, every three days, or daily) and isolation of asymptomatic persons compared to symptom-based testing and isolation | USA | Nursing homes | Residents and staff |
| Shirreff | Measuring basic reproduction number to assess effects of nonpharmaceutical interventions on nosocomial SARS-CoV-2 transmission | July 2022 | Emerging Infectious Diseases | Stochastic Compartmental | Estimate R0 and patient-to-patient transmission in various wards of a hospital, following changes in testing capacity | France | Hospital | Patients |
| Smith | Optimizing COVID-19 surveillance in long-term care facilities: a modelling study | Dec 2020 | BMC Infectious Diseases | Stochastic Agent-based | Explore various testing strategies for detecting outbreaks in LTCFs | France | LTCF | Residents and staff |
| Smith | Rapid antigen testing as a reactive response to surges in nosocomial SARS-CoV-2 outbreak risk | Jan 2022 | Nature Communications | Stochastic Agent-based | Estimate health and economic benefits of using rapid antigen testing to control outbreaks in LTCFs | France | LTCF | Residents and staff |
| Stevenson | Modelling of hypothetical SARS-CoV-2 point-of-care tests on admission to hospital from A&E: rapid cost-effectiveness analysis | Mar 2021 | Health Technology Assessment | Stochastic Agent-based | Estimate effectiveness and cost-effectiveness of point-of-care tests compared to laboratory-based tests. Assess occupancy levels in hospital areas such as waiting bays | UK (NHS) | Hospital | Patients and HCWs |
| Stevenson | Modelling of hypothetical SARS-CoV-2 point of care tests for routine testing in residential care homes: rapid cost-effectiveness analysis. | June 2021 | Health Technology Assessment | Stochastic Agent-based | Cost-effectiveness of testing strategies | UK (NHS) | Care home | Residents and staff |
| Tofighi | Modelling COVID -19 Transmission in a Hemodialysis Centre Using Simulation Generated Contacts Matrices | Nov 2021 | PLoS ONE | Stochastic Agent-based | Estimate contact matrices, explore outbreak scenarios and evalute testing and cancelling breaks | Canada | Dialysis units | Patients and HCWs |
| Tsoungui Obama | Preventing COVID-19 spread in closed facilities by regular testing of employees—An efficient intervention in long-term care facilities and prisons? | Apr 2021 | PLoS ONE | Deterministic Compartmental | Assess the efficacy of regular testing in long-term care facilities | Germany | LTCF (nursing homes, prisons) | Residents and staff |
| Vilches | Multifaceted strategies for the control of COVID-19 outbreaks in longterm care facilities in Ontario, Canada | July 2021 | Preventive Medicine | Stochastic Agent-based | Assess the impact of PPE, routine testing of staff and vaccination of staff and residents | Canada | LTCF, nursing homes | Residents and staff |
| Wilmink | Real-Time Digital Contact Tracing: Development of a System to Control COVID-19 Outbreaks in Nursing Homes and Long-Term Care Facilities | Aug 2020 | JMIR Public Health Surveillance | Deterministic Compartmental | - development and implementation of a real-time digital contact tracing system designed specifically for LTCFs - assess the number of cases and resultant deaths for several intervention types: digital or manual contact tracing, symptom-based mapping, PCR testing, and no intervention | USA | LTCFs including nursing homes | Residents |
| Yuval | Optimizing testing policies for detecting COVID-19 outbreaks | July 2020 | Arxiv | Stochastic Compartmental | Study the possibility for nursing homes to utilize a fixed test budget to detect oubreaks | / | Nursing homes | Residents and staff with no distinction |
| Zhang | Evaluating the Need for Routine COVID-19 Testing of Emergency Department Staff: Quantitative Analysis | Dec 2020 | JMIR Public Health Surveillance | Stochastic Compartmental | Assessing the benefits of periodic testing of HCWs | United States | Emergency departments | Patients and HCWs |
| Zhou | Agent-Based Simulation of Virus Testing in Certain-Exposure Time through Community Health Service Centers' Evaluation-A Case Study of Wuhan. | Nov 2021 | Healthcare | Stochastic Agent-based | Performance of community testing facilities in Wuhan, with respect to speed and completeness at which the local population is tested | Wuhan | Testing room in community health service centres | Community members tested |

### PRISMA 2020 Main Checklist

| **Topic** | **No.** | **Item** | **Location where item is reported** |
| --- | --- | --- | --- |
| **TITLE** |  |  |  |
| **Title** | 1 | Identify the report as a systematic review. | Line 1 |
| **ABSTRACT** |  |  |  |
| **Abstract** | 2 | See the PRISMA 2020 for Abstracts checklist |  |
| **INTRODUCTION** |  |  |  |
| **Rationale** | 3 | Describe the rationale for the review in the context of existing knowledge. | Lines 68-74 |
| **Objectives** | 4 | Provide an explicit statement of the objective(s) or question(s) the review addresses. | Lines 74-78 |
| **METHODS** |  |  |  |
| **Eligibility criteria** | 5 | Specify the inclusion and exclusion criteria for the review and how studies were grouped for the syntheses. | Supplementary file, section 2 |
| **Information sources** | 6 | Specify all databases, registers, websites, organisations, reference lists and other sources searched or consulted to identify studies. Specify the date when each source was last searched or consulted. | Supplementary file, section 1 |
| **Search strategy** | 7 | Present the full search strategies for all databases, registers and websites, including any filters and limits used. | Supplementary file, section 1 |
| **Selection process** | 8 | Specify the methods used to decide whether a study met the inclusion criteria of the review, including how many reviewers screened each record and each report retrieved, whether they worked independently, and if applicable, details of automation tools used in the process. | Supplementary file, section 3 |
| **Data collection process** | 9 | Specify the methods used to collect data from reports, including how many reviewers collected data from each report, whether they worked independently, any processes for obtaining or confirming data from study investigators, and if applicable, details of automation tools used in the process. | Supplementary file, section 4 |
| **Data items** | 10a | List and define all outcomes for which data were sought. Specify whether all results that were compatible with each outcome domain in each study were sought (e.g. for all measures, time points, analyses), and if not, the methods used to decide which results to collect. | Supplementary file, section 4 |
|  | 10b | List and define all other variables for which data were sought (e.g. participant and intervention characteristics, funding sources). Describe any assumptions made about any missing or unclear information. | Supplementary file, section 4 |
| **Study risk of bias assessment** | 11 | Specify the methods used to assess risk of bias in the included studies, including details of the tool(s) used, how many reviewers assessed each study and whether they worked independently, and if applicable, details of automation tools used in the process. | Not applicable |
| **Effect measures** | 12 | Specify for each outcome the effect measure(s) (e.g. risk ratio, mean difference) used in the synthesis or presentation of results. | Not applicable |
| **Synthesis methods** | 13a | Describe the processes used to decide which studies were eligible for each synthesis (e.g. tabulating the study intervention characteristics and comparing against the planned groups for each synthesis (item 5)). | Not applicable |
|  | 13b | Describe any methods required to prepare the data for presentation or synthesis, such as handling of missing summary statistics, or data conversions. | None |
|  | 13c | Describe any methods used to tabulate or visually display results of individual studies and syntheses. | Not applicable |
|  | 13d | Describe any methods used to synthesize results and provide a rationale for the choice(s). If meta-analysis was performed, describe the model(s), method(s) to identify the presence and extent of statistical heterogeneity, and software package(s) used. | Not applicable |
|  | 13e | Describe any methods used to explore possible causes of heterogeneity among study results (e.g. subgroup analysis, meta-regression). | Not applicable |
|  | 13f | Describe any sensitivity analyses conducted to assess robustness of the synthesized results. | Not applicable |
| **Reporting bias assessment** | 14 | Describe any methods used to assess risk of bias due to missing results in a synthesis (arising from reporting biases). | Not applicable |
| **Certainty assessment** | 15 | Describe any methods used to assess certainty (or confidence) in the body of evidence for an outcome. | Not applicable |
| **RESULTS** |  |  |  |
| **Study selection** | 16a | Describe the results of the search and selection process, from the number of records identified in the search to the number of studies included in the review, ideally using a flow diagram. | Lines 87-88 + Supplementary figure S1 |
|  | 16b | Cite studies that might appear to meet the inclusion criteria, but which were excluded, and explain why they were excluded. | Supplementary figure S1 |
| **Study characteristics** | 17 | Cite each included study and present its characteristics. | Supplementary Table |
| **Risk of bias in studies** | 18 | Present assessments of risk of bias for each included study. | Not applicable |
| **Results of individual studies** | 19 | For all outcomes, present, for each study: (a) summary statistics for each group (where appropriate) and (b) an effect estimate and its precision (e.g. confidence/credible interval), ideally using structured tables or plots. | Not applicable |
| **Results of syntheses** | 20a | For each synthesis, briefly summarise the characteristics and risk of bias among contributing studies. | Not applicable |
|  | 20b | Present results of all statistical syntheses conducted. If meta-analysis was done, present for each the summary estimate and its precision (e.g. confidence/credible interval) and measures of statistical heterogeneity. If comparing groups, describe the direction of the effect. | Lines 88-100 + Figure 2 |
|  | 20c | Present results of all investigations of possible causes of heterogeneity among study results. | Not applicable |
|  | 20d | Present results of all sensitivity analyses conducted to assess the robustness of the synthesized results. | Not applicable |
| **Reporting biases** | 21 | Present assessments of risk of bias due to missing results (arising from reporting biases) for each synthesis assessed. | Not applicable |
| **Certainty of evidence** | 22 | Present assessments of certainty (or confidence) in the body of evidence for each outcome assessed. | Not applicable |
| **DISCUSSION** |  |  |  |
| **Discussion** | 23a | Provide a general interpretation of the results in the context of other evidence. | Lines 238-246 |
|  | 23b | Discuss any limitations of the evidence included in the review. | Lines 247-283 + 287-296 |
|  | 23c | Discuss any limitations of the review processes used. | Lines 319-328 |
|  | 23d | Discuss implications of the results for practice, policy, and future research. | Lines 283-286 + 297-318 |
| **OTHER INFORMATION** |  |  |  |
| **Registration and protocol** | 24a | Provide registration information for the review, including register name and registration number, or state that the review was not registered. | Not applicable |
|  | 24b | Indicate where the review protocol can be accessed, or state that a protocol was not prepared. | Not applicable |
|  | 24c | Describe and explain any amendments to information provided at registration or in the protocol. | Not applicable |
| **Support** | 25 | Describe sources of financial or non-financial support for the review, and the role of the funders or sponsors in the review. | Line 340- |
| **Competing interests** | 26 | Declare any competing interests of review authors. | Line 338- |
| **Availability of data, code and other materials** | 27 | Report which of the following are publicly available and where they can be found: template data collection forms; data extracted from included studies; data used for all analyses; analytic code; any other materials used in the review. | Line 88 |

### PRIMSA Abstract Checklist

| **Topic** | **No.** | **Item** | **Reported?** |
| --- | --- | --- | --- |
| **TITLE** |  |  |  |
| **Title** | 1 | Identify the report as a systematic review. | Yes |
| **BACKGROUND** |  |  |  |
| **Objectives** | 2 | Provide an explicit statement of the main objective(s) or question(s) the review addresses. | Yes |
| **METHODS** |  |  |  |
| **Eligibility criteria** | 3 | Specify the inclusion and exclusion criteria for the review. | Yes |
| **Information sources** | 4 | Specify the information sources (e.g. databases, registers) used to identify studies and the date when each was last searched. | Yes |
| **Risk of bias** | 5 | Specify the methods used to assess risk of bias in the included studies. | No |
| **Synthesis of results** | 6 | Specify the methods used to present and synthesize results. | Yes |
| **RESULTS** |  |  |  |
| **Included studies** | 7 | Give the total number of included studies and participants and summarise relevant characteristics of studies. | No |
| **Synthesis of results** | 8 | Present results for main outcomes, preferably indicating the number of included studies and participants for each. If meta-analysis was done, report the summary estimate and confidence/credible interval. If comparing groups, indicate the direction of the effect (i.e. which group is favoured). | Yes |
| **DISCUSSION** |  |  |  |
| **Limitations of evidence** | 9 | Provide a brief summary of the limitations of the evidence included in the review (e.g. study risk of bias, inconsistency and imprecision). | No |
| **Interpretation** | 10 | Provide a general interpretation of the results and important implications. | Yes |
| **OTHER** |  |  |  |
| **Funding** | 11 | Specify the primary source of funding for the review. | No |
| **Registration** | 12 | Provide the register name and registration number. | No |

*From:* Page MJ, McKenzie JE, Bossuyt PM, Boutron I, Hoffmann TC, Mulrow CD, et al. The PRISMA 2020 statement: an updated guideline for reporting systematic reviews. MetaArXiv. 2020, September 14. DOI: 10.31222/osf.io/v7gm2. For more information, visit: www.prisma-statement.org
